# Variable-Interval Temporal Feathering to Optimize Organ-at-Risk Repair for Head and Neck Adaptive Radiotherapy

**DOI:** 10.1101/2024.11.07.24316948

**Authors:** Aysenur Karagoz, Mehdi Hemmati, Fatemeh Nosrat, Panayiotis Mavroidis, Cem Dede, Lucas B. McCullum, Raul Garcia, Seyedmohammadhossein Hosseinian, Jacob G. Scott, James E. Bates, Heiko Enderling, Abdallah S.R. Mohamed, Kristy K. Brock, Andrew J. Schaefer, Clifton D. Fuller, Rice/MD Anderson Center for Operations Research in Cancer (CORC) and MD Anderson Head and Neck Cancer Symptom Working Group.

**Author notes:** **Co-Corresponding Authors:** Mehdi Hemmati, PhD; Andrew J. Schaefer, PhD; Clifton D. Fuller, MD, PhD. **Author Responsible for Statistical Analysis:** Mehdi Hemmati, PhD. **Data Availability Statement:** All data generated and analyzed during this study are included in this article (and its supplementary document). The source code is accessible through https://figshare.com/s/adc2470447ea492338c2. **CRediT Statement:** Conceptualization: **AK, MH, PM, CD, RG, SH, JGS, JEB, HE, ASRM, KKB, AJS, CDF**; Methodology: **AK, MH, RG, LBM, PM, CD, SH, AJS, CDF**; Software: **AK, MH**; Validation: **AK, MH**; Formal Analysis: **AK, MH, PM, SH**; Investigation: **AK, MH, RG, JGS, CD, AJS, CDF**; Resources: **CD, JGS, ASRM, AJS, CDF**; Data Curation: **AK, MH, CD, ASRM**; Writing - Original Draft: **AK, MH**; Writing - Review & Editing: **AK, MH, FN, PM, CD, LBM, RG, SH, JGS, JEB, HE, ASRM, KKB, AJS, CDF**; Visualization: **AK, MH**; Supervision: **JGS, ASRM, AJS, CDF**; Project Administration: **AJS, CDF**; Funding Acquisition: **AJS, CDF**.

## Abstract

**Purpose:** Temporally feathered radiation therapy (TFRT) for head-and-neck cancer (HNC) radiotherapy combines variable-dose daily subplans to increase the rest time of organs-at-risk (OARs) as sought in intensity modulated radiation therapy (IMRT). While the standard TFRT recommends uniform rest time for each OAR, improved toxicity outcomes may be achieved through variable rest time for OARs by incorporating the OARs’ variable radiosensitivity profiles.

**Methods and Materials:** A decision-making model was constructed to maximize the combined recovery of OARs by determining OARs’ optimal rest times. Two main components were incorporated: the cumulative biologically effective dose based on the linear-quadratic model; and a dynamical model capturing the adjusted recovery of OARs as a function of delivered dose. Further, variable radiosensitivity profiles were allowed across the OARs to capture their variable recovery time. Individual recoveries of each OAR under IMRT and the standard TFRT (sTFRT) was compared against optimized TFRT (oTFRT).

**Results:** Five OARs (larynx, esophagus, parotid, spinal cord, brainstem) were considered. When the cumulative dose delivered under TFRT and IMRT remains the same, three OARs exhibited higher recovery under oTFRT compared to the second-best approach (larynx (81.8% vs. 74.1%), esophagus (95.9% vs. 93.9%), parotid (85.6% vs. 83.5%), while the recovery of spinal cord (90.5% vs. 90.8%) and brainstem (96.2% vs. 96.6%) remained comparable under TFRT and IMRT approaches. With different cumulative dose under TFRT and IMRT, oTFRT achieved significantly higher recovery for larynx (95.5% vs. 81.8%) and parotid (92.9% vs. 85.6%), while it is slightly outperformed by IMRT for esophagus (93.4% vs. 95.9%), spinal cord (87.1% vs. 90.5%), and brainstem (90.2% vs. 96.6%). When considering the minimum end-of-treatment recovery, oTFRT always achieved higher recovery among the other two approaches.

**Conclusions:** By considering non-identical radiosensitivity profiles of OARs in HNC radiotherapy, TFRT can optimize their rest time to enhance recovery at the end of treatment, potentially reducing patient toxicities.

## Introduction

More than 71,000 Americans will be afflicted by head-and-neck cancer (HNC) in 2024, accounting for about 4% of all US cancer cases.^1^ Intensity-modulated radiation therapy (IMRT) in combination with cisplatin chemotherapy remains the primary curative treatment approach for HNC patients.^2,3,4^ IMRT has been successful in terms of oncological results, leading to increased locoregional control and survival in HNC patients^5^. With the increasing predominance of human papillomavirus (HPV)-related cases, which have a significantly better prognosis compared to HPV-negative cases (80% five-year survival rate for HPV-positive cases vs. 40% for HPV-negative cases^6^), the five-year overall survival rate of HNC is now 68.5%.^7^

The majority of HNC patients treated with IMRT, however, suffer from both acute and chronic toxicities including xerostomia, dysphagia, and osteoradionecrosis, leading to a significant decline in their quality of life. These sequelae stem directly from the injury to the non-target organs-at-risk (OARs) while delivering high-dose radiation to the tumor.^8^ With an improved survival rate and a focal shift of radiation therapy (RT) treatment planning toward sparing normal tissues, improving the quality of life in HNC survivors has now become a primary goal in HNC radiation therapy treatment planning.^9^

The current standard IMRT technique for HNC prescribes a fixed daily dose fraction of (approximately) 2 Gy to the tumor, consequently imposing a fixed daily dose to surrounding OARs. This is consistent with the premise that a fractionated plan based on a fixed daily dose to OARs allows enough time for each OAR recovery between treatment sessions. However, there has been a long-lasting debate in the oncology community, questioning the optimality of daily fractions of equal doses in radiation therapy. Recent studies report potential benefits of variable fraction dose based on the non-homogeneity observed in the tumor cells and late-treatment effects such as increased tumor repopulation rate.^10^ Nonuniform fractionation is also an artifact of adaptive radiation therapy (ART) – the technique that advocates adaptation of the treatment plan as a function of tumor/OAR responses, which can potentially lead to variable daily doses. Regardless of the treatment planning technique, most studies that advocate nonuniform fractionation focus on hypofractionating subregions of the tumor while delivering a uniform, daily fixed dose to healthy tissues.^11,12^

Recently, Lopez Alfonso et al.^13^ proposed a novel RT treatment planning strategy, referred to as temporally feathered RT (TFRT), that leverages time to allow variable daily doses to OARs with the aim of improving sublethal damage repair. A TFRT plan is constructed as a composite of several subplans, each delivered on a specific weekday. The number of OARs varies between cancer sites; for HNC patients, five OARs are typically considered.^13^ Each weekday, the designated OAR is allowed to receive a slightly higher amount of dose (but remains constrained subject to a maximum dose threshold to avoid OAR lethal damage), enabling the dose-optimization algorithm to reduce radiation to all other OARs on that day. (The OARs receiving a slightly higher amount of dose are referred to as the “temporally feathered” OARs for that day.^13^) Therefore, each OAR is administered a higher-than-standard fractional dose of IMRT once weekly, followed by lower fractional doses in the remaining four days. The hypothesis is that the one-week interval between the higher fractional doses will promote sublethal damage repair and repopulation, which are shown to take weeks to months to occur after the peak acute toxicity.^14^ Simulated and in vivo studies indicate the success of TFRT in reducing toxicity burden for HNC patients when the higher-than-standard doses are carefully chosen.^15,16^

The current TFRT framework computes the composite plan based on the premise that all OARs share a similar response to the delivered dose, thus uniformly allowing a one-week recovery between two consecutive high-dose days for each OAR. Using the well-known linear-quadratic model,^17^ this is equivalent to having the same radiosensitivity parameters for all OARs in HNC radiation therapy. However, in practice, OARs exhibit different levels of radiosensitivity, thus some OARs may have higher recovery rates while others require longer rest time. For example, the brainstem and spinal cord (two common OARs in HNC radiation therapy) exhibit significantly different levels of radiosensitivity: the brainstem is more sensitive to radiation dose and thus gains relatively higher benefit from fractionation.^18^ A relevant question regards the feasibility of TFRT when the differences in radiosensitivity profiles are incorporated to allow nonuniform recovery time between two high-dose days, and whether the nonuniform recovery time may lead to lower radiation-induced toxicity in the organ.

As our first effort to present the concept of a variable schedule for OAR feathering, this study investigates the effect of variable radiosensitivity profiles across OARs on the outcomes of TFRT. Specifically, we will develop a mathematical optimization model that allows variable recovery time between high-dose days by computing the optimal “OAR feathering” scheme for TFRT treatment planning. Further, we leverage our model to study the outcomes of TFRT in the presence of specific organ-preserving goals, which can be set according to the clinician’s preferences.

## Methods and Materials

We develop a nonlinear mixed-integer optimization model that computes the optimal OAR feathering schedule with the aim of maximizing organ recovery by the end of the treatment. In constructing our model, we utilize two main biological concepts, namely the linear-quadratic model^17^ and the non-spatial dynamical model of tissue response to radiation,^19^ to incorporate the fraction of OAR cells surviving the radiation and the recovery/repair in the damaged healthy tissues, respectively.

### The linear-quadratic model

The linear-quadratic (LQ) model^17^ predicts the cell survival likelihood as a function of radiation dose *d* (Gy), given by

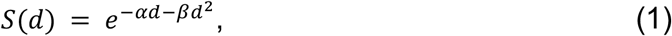

where *α* (Gy^-1^) and *β* (Gy^-2^) are tissue-specific radiosensitivity parameters. The ratio *α*/*β* is used to measure the tissue’s sensitivity to fraction dose: while tumor cells tend to exhibit higher values of *α*/*β*, the healthy tissues often have lower values of *α*/*β*, making them more sensitive to fractionation.^20^

For *n* fractions each delivering *d* Gys, the biologically effective dose (BED)^21^ is defined as −*ln*(*S*)/*α*, where *S* is defined according to the LQ model (1), i.e.,

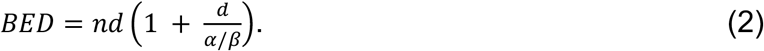

When only uniform treatment plans are considered, the BED equation (2) is utilized to evaluate and compare the biological effect of two or more radiation therapy treatments, by accounting for the changes in dose-per-fraction, total dose, and the number of fractions.

In contrast, when the daily dose received by an OAR is nonuniform across treatment days (as is the case in TFRT treatment planning), the BED equation (2) becomes

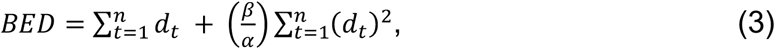

where *d*_*t*_ is the dose delivered to OAR in fraction *t* = 1, . . ., *n*. We will utilize the BED equation (3) to incorporate the nonuniform dose received by OARs in a TFRT treatment plan while allowing OAR-specific *α*/*β* ratios.

### Cells recovery from radiation-induced damage

The ultimate goal in deriving the optimal OAR feathering schedule is to minimize the toxicities incurred by the OARs. In the literature, this is commonly modeled as a normal tissue complication probability (NTCP) model. Specifically, NTCP models calculate the likelihood of OAR damage, manifested through radiation-induced side effects. Several NTCP models have been proposed in the last two decades, ranging from purely dose-based mechanistic models^22,23,24,25^ to more recent data-driven models.^26,27,28,29^

In this work, we will adopt the nonspatial dynamical model of tissue response to radiation by Hanin and Zaider,^19^ which was originally used to evaluate the benefits of TFRT in reducing the OARs toxicities.^13^ We let 0 ≤ *N*(*t*) ≤ 1 represent the level of normal tissue recovery (with small values of *N*(*t*) indicating severe damage). Then, the cell recovery process can be represented by the following ordinary differential equation (ODE):

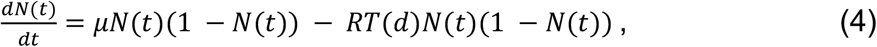

where *μ* is the organ-specific parameter representing the recovery rate and *RT*(*d*) represents the likelihood of damaging each cell of the OAR receiving a radiation dose *d*. Using the LQ model (1), *RT*(*d*) = 1 − *S*(*d*), i.e., *RT*(*d*) represents the damaged fraction of the OAR when receiving a (single-fraction) radiation dose *d*.

The recovery model (4) is a form of NTCP model as it represents a quantitative measure of radiation-induced damage to an OAR.^13^ Our choice of this recovery model is further motivated by the presence of both *α* and *β* parameters (in the definition of *RT*(*d*)), which relates cell recovery to the radiosensitivity of the OAR, similar to the BED equation (3).

Mathematical optimization models that incorporate ODE similar to (4) often present computational challenges. To approximate (4), Runge-Kutta (RK) methods^30^ can be used. Here, we utilize the (forward) Euler’s method, which is the first-order RK method, previously applied in chemotherapy optimization^31^. We approximate the ODE (4) by a set of difference equations, as follows:

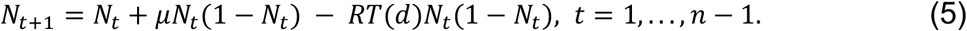

Our optimization model directly utilizes Equation (5) as a measure to guide decisions on OAR feathering in TFRT treatment planning.

### Mathematical optimization model

We will develop a mathematical optimization problem which derives an optimal OAR feathering schedule and the optimal (possibly nonuniform) daily dose received by each OAR while maintaining the total dose to OARs (as is prescribed in the standard IMRT), to maximize the OARs recovery by the end of treatment. We define *M* and *n* as the number of OARs and the number of fractions (treatment days), respectively.

#### Constraints on maximum BED received by the OARs

While TFRT allows a slightly higher dose to a designated OAR each weekday, the BED of the dose received by the OAR still cannot become excessive. We let the parameters *τ*_*m*_, *m* = 1, . . ., *M* be the maximum allowable BED received by the OAR *m* by the end of the treatment. Given *m* = 1, . . ., *M* and *t* = 1, . . ., *n*, we define the decision variable 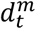 as the dose received by OAR *m* in fraction *t*, and enforce the following constraint

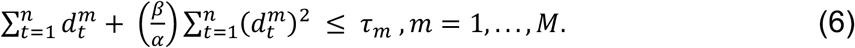

#### Constraints on daily subplan selection

TFRT requires computing several treatment subplans, one for each day when an OAR is feathered (i.e., will receive a daily dose higher than the standard IMRT dose, while other OARs are allowed to receive a lower radiation dose compared to the standard IMRT dose).

If the number of OARs in TFRT treatment planning is *M*, then this necessitates the inclusion of *M* (standard IMRT-based) dose-optimization subproblems, each corresponding to a weekday, within the TFRT treatment planning procedure. Since solving the dose-optimization subproblem is computationally challenging, the resulting TFRT treatment planning optimization problem becomes significantly more difficult to solve.

To address this issue, we will leverage one of the key assumptions in TFRT, which maintains a fixed daily dose to the tumor. We further utilize the notion of sparing factor,^32,33,34^ which allows computation of the dose received by each OAR as a function of the dose delivered to the tumor. Specifically, if the sparing factor of OAR *m* in the treatment subplan for fraction *t* is denoted by 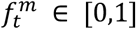, then the dose delivered to the OAR in fraction *t* is computed as 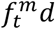, where *d* is the daily (fixed) dose delivered to the tumor. Accordingly, each treatment subplan comprises *M*sparing factors, one corresponding to each OAR. By varying an OAR’s sparing factors across treatment days, we allow a nonuniform daily dose to be received by the OAR.

For each subplan *k* = 1, . . ., *M*, we denote by *p*^*k*^ an *M*-dimensional vector, comprising individual OAR’s sparing factors *p*^*k,m*^ ∈ [0,1], *m* = 1, . . ., *M*. Accordingly, we define binary decision variables 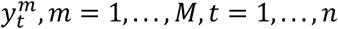, taking the value of one if the subplan corresponding to the designated OAR *m* is administered in fraction *t*. We enforce

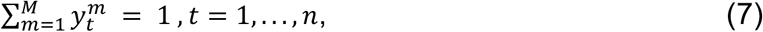

and

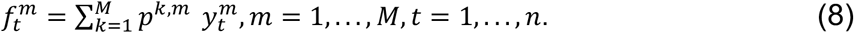

Equation (7) implies that only one subplan is selected for each weekday, indicating exactly one OAR is feathered. The definition of sparing factor of an OAR in any fraction is enforced through Equation (8). Since the daily dose to the tumor is 2 Gy, we enforce

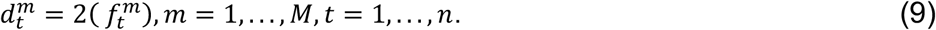

#### Constraints on OAR feathering based on its recovery

In our model, the recovery of an OAR *m* is computed as in Equation (5), i.e.,

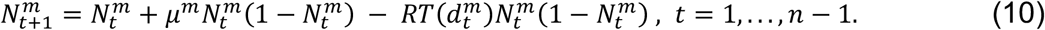

Similar to the original study of TFRT, we assume that the OAR is at tissue homeostasis before radiation with a 1% turnover rate^13^, hence we enforce the initial following initial condition

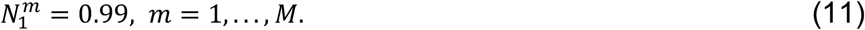

We will leverage the recovery level of each OAR to guide our choice of the subplan to be administered each weekday, i.e., the value of the decision variables 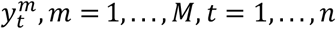. We define *ξ*^*m*^ as the threshold for the minimum recovery level required for OAR *m* = 1, . . ., *M* before it can receive a dose higher than the daily standard IMRT dose. We enforce

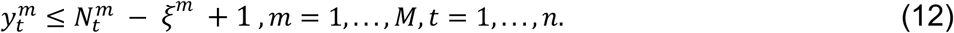

#### Integrated Model

We define *w*^*m*^ as the weight (indicating the importance) of the OAR *m* = 1, . . ., *M*. Our model is presented as follows

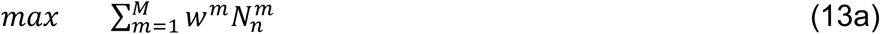

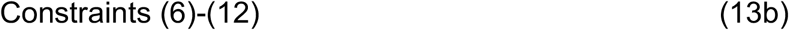

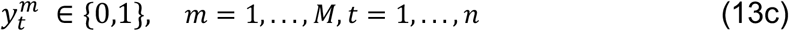

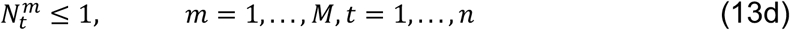

The proposed model (13) seeks the maximum weighted OARs recovery at the end of the treatment subject to the constraints introduced earlier in this section. Constraints (13c) imply the definition of the *y* decision variables and constraints (13d) enforce the logical upper bound on the recovery of each OAR. We note that our choice of the objective function (13a) allows the inclusion of the clinician’s preference in sparing certain OARs by considering larger weights in the objective function. Accordingly, we consider two classes of model (13) in our numerical study, 1) when the OARs have the same weight, and 2) when the recovery of certain OARs may have a higher priority, thus resulting in nonuniform weights across OARs.

Note that Model (13) does not require identical *total* dose delivery under IMRT and TFRT. In fact, as reported in the study by Alfonso et al.^13^, the total cumulative dose received by OARs under TFRT may be slightly more than IMRT. Hence, we will also consider two variations of Model (13) depending on whether or not the total dose received by OARs under TFRT is at least the total dose delivered under IMRT. To enforce this constraint, we define 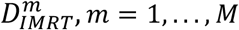 be the total dose received by each OAR in standard IMRT. We impose

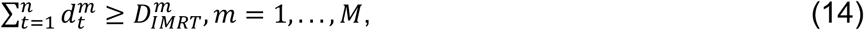

to Model (13). To distinguish the two cases, we refer to this model as “Dose-restricted TFRT” and refer to Model (13) (without Constraints (14)) as “Dose-free TFRT”.

Finally, we note that Model (13) includes an exponential term within constraint (10), which poses computational challenges. Thus, we conducted experiments substituting the exponential term with its quadratic midpoint approximation to facilitate problem-solving. Our analysis focused on five OARs, each characterized by distinct radiosensitivity and recovery parameters.

#### Data

To illustrate the advantages of using our proposed model over standard TFRT treatment planning, we develop the following numerical experiment comprising five OARs for feathering, inspired by the parameters reported in the first TFRT study^13^. Notably, Alfonso et al.^13^ apply a fixed *α*/*β* ratio of 3 across the five OARs considered in their study with *α* = 0.3 and *β* = 0.1. To recognize the presence of OARs with different dose-response profiles, we allow variable radiosensitivity parameters in our OARs by scaling the values of *α* and *β* reported by Kehwar^33^ for the brainstem, larynx, esophagus, parotid, and spinal cord; the scaling is performed to ensure using relatively comparable magnitudes for *α* and *β*, as were used in the original TFRT study.^13^ Table 1 reports the radiosensitivity parameters chosen for this experiment. We note that our chosen *α*/*β* ratio values are consistent with those in the literature (which varies between 2 and 4 for late-responding OARs). We have further scaled down the recovery rates reported in the TFRT study, and have used a universal recovery threshold, *ξ* = 0.75, which indicates that an OAR can not be feathered if its recovery level falls below 0.75. Finally, we have obtained the maximum allowable values for the total BED received by each OAR based on ^36,37^. See Table 1.

**Table 1:** Radiosensitivity parameters (*α, β*), recovery rate (*μ*), recovery threshold (*ξ*), and maximum dose allowed to the OAR (*τ*). Lopez et al.^**13**^ considered α values of 0.15, 0.21 and 0.3. We adjusted the values in the literature^**35**^ accordingly to ensure comparability and different radiosensitivity for OARs.

| OARs | $\alpha$ (Gy<sup>-1</sup>)<sup>13,34</sup> | $\beta$ (Gy<sup>-2</sup>)<sup>13,35</sup> | $\mu$ (day<sup>-1</sup>) | $\xi$ | $\tau$ (Gy)<sup>36,37</sup> |
| --- | --- | --- | --- | --- | --- |
| Brainstem | 0.1964 | 0.0936 | 0.095 | 0.75 | 95 |
| Larynx | 0.3252 | 0.0836 | 0.05 | 0.75 | 110 |
| Esophagus | 0.2832 | 0.0944 | 0.06 | 0.75 | 92 |
| Parotid | 0.2512 | 0.0836 | 0.075 | 0.75 | 117 |
| Spinal Cord | 0.178 | 0.054 | 0.09 | 0.75 | 104 |

For the sparing factor, we used the IMRT plan generated by the TFRT treatment planning guidelines proposed by Parsai et al.^15^. For each OAR, the base sparing factor is computed as the ratio of the (fixed) daily dose received by the OAR and the fixed daily dose delivered to the tumor, i.e., 2 Gy. Next, one subplan corresponding to each OAR is constructed by fixing the sparing factor of the to-be-feathered OAR at 1, and scaling down the sparing factors for the remaining OARs by 0.25 or 0.45. Table 2 reports each of the TFRT subplans and the corresponding sparing factors.

**Table 2:** Sparing factors for the experiments were computed using the daily dose received by the OARs.^**15**^ The base sparing factor was calculated as the ratio of the fixed daily dose received by the OAR to the daily dose delivered to the tumor (2 Gy). For each OAR, a subplan is created by setting the sparing factor of the targeted OAR to 1 and scaling down the factors for the other OARs by 0.25 or 0.45.

| OARs | Sparing factors<sup>15</sup> |  |  |  |  |  | Total dose received by IMRT (Gy)<sup>15</sup> |
| --- | --- | --- | --- | --- | --- | --- | --- |
|  | Plan 1 | Plan 2 | Plan 3 | Plan 4 | Plan 5 | IMRT |  |
| Brainstem | 1.0 | 0.0951 | 0.2151 | 0.0335 | 0.1551 | 0.3 | 21.0 |
| Larynx | 0.092 | 1.0 | 0.042 | 0.1045 | 0.0295 | 0.25 | 17.5 |
| Esophagus | 0.005 | 0.005 | 1.0 | 0.005 | 0.005 | 0.204 | 14.28 |
| Parotid | 0.18 | 0.1164 | 0.2436 | 1.0 | 0.0528 | 0.318 | 22.26 |
| Spinal Cord | 0.1418 | 0.3884 | 0.2651 | 0.2651 | 1.0 | 0.411 | 28.77 |

#### Performance metrics

To compare the outcomes of standard IMRT versus our optimized TFRT with variable feathering schedule, we compare 1) the end-of-treatment recovery level and 2) the dose delivered for each OAR. In addition to the standard IMRT and our optimized TFRT schedule, we also consider the standard TFRT of Alfonso et al.^13^, referred to as the “standard TFRT” to indicate that it feathers each OAR on a specific weekday, known a priori, with a standard one-week recovery period between high-dose days.

## Results

In the base analysis, we implemented the dose-restricted TFRT (Model (13) coupled with Constraints (14)) and dose-free TFRT (Model (13) alone), while assuming the OAR weights (*w*^*m*^) in the TFRT model are identical. We further considered a hypothetical case in which the clinician may have a preference in prioritizing the recovery of certain OAR (e.g., spinal cord) over other OARs. In this case, the clinician’s preference can be modeled by setting the weights (**Supplementary Document A**).

### Dose-restricted TFRT with identical recovery weights

Figure 1. shows the result of our base analysis for dose-restricted TFRT. According to **Figure 1a**, Each OAR was feathered exactly seven times throughout the treatment to match the cumulative dose delivered under the optimized TFRT with that delivered in standard IMRT. Brainstem and spinal cord were feathered more frequently during the first 15 fractions. The other OARs were feathered on the remaining days with no more than two consecutive days of feathering. On high-dose days, OARs received a dose of 2 Gy, while on rest days, they were administered less than 1 Gy. Considering its highest sparing factor, the spinal cord received the highest dose on the lower-dose days. In contrast, the esophagus received minimal doses on these days due to its low sparing factor of 0.005. As indicated in **Figure 1b**, significant decrease in each OAR’s recovery was observed during feathering periods and a slight increase when lower doses were administered. Throughout the treatment, none of the OARs fell below the recovery threshold of *ξ* = 0.75. The recovery levels at the end of the treatment for the brainstem, larynx, esophagus, parotid and spinal cord were observed as 96.2%, 81.8%, 95.9%, 85.6% and 90.5%, respectively.

**Figure 1:**
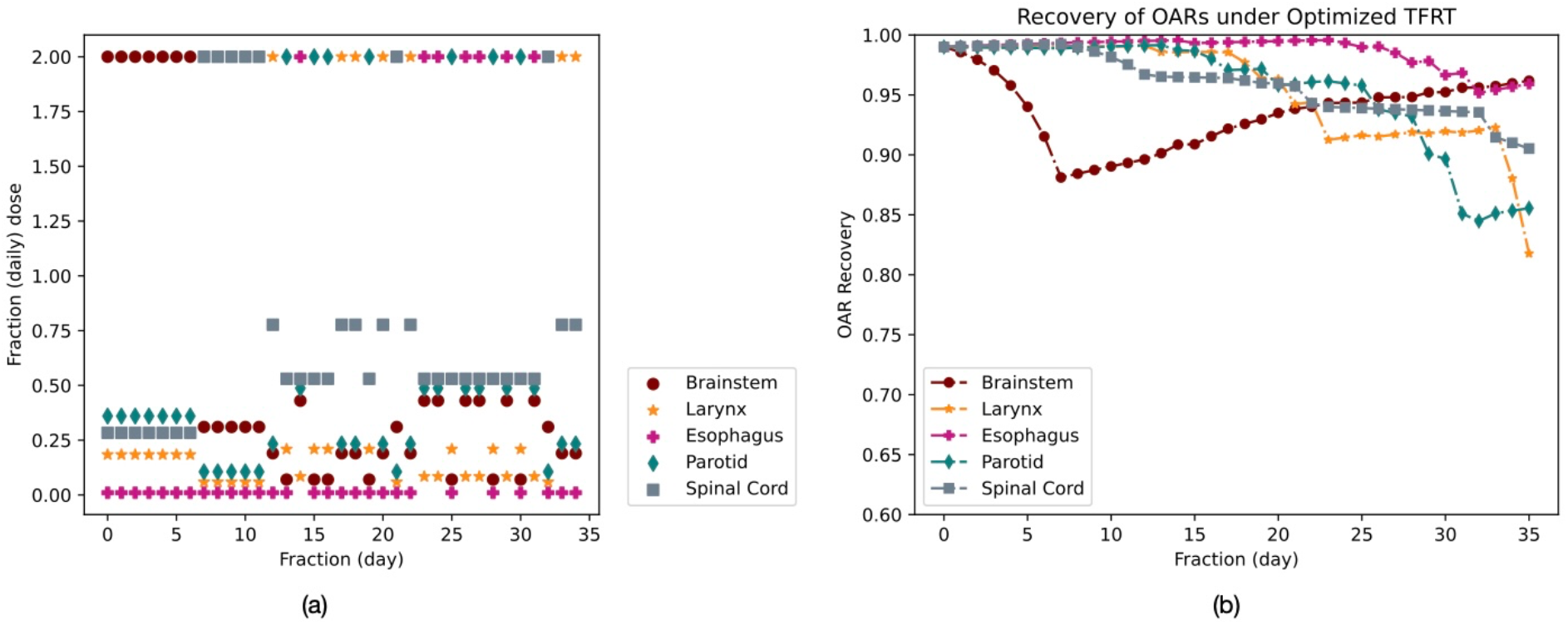
(a) The feathering schedule and (b) the recovery of OARs under optimized dose-restricted TFRT; OAR: organ-at-risk, TFRT: temporally feathered radiation therapy.

**Figure 2** shows, for each OAR, the individual recovery and the cumulative dose received under each of the three treatment techniques, namely the standard IMRT, the standard TFRT, and the optimized TFRT. While the end-of-treatment cumulative dose in this analysis remained identical among the three approaches (**Figures 2a-2e**), the on-treatment cumulative dose delivered under TFRT-based approached to all OARs except brainstem remained lower than that delivered under the standard IMRT. **Figure 2f** illustrates the recovery of the brainstem across these treatment approaches. Given that the brainstem has the lowest *α*/*β* ratio and relatively smaller *α* and *β* values, it exhibits better recovery when exposed to lower fractional doses. Consequently, by the end of treatment, the brainstem’s recovery under optimized TFRT (96.2%) was comparable to both standard TFRT (96.3%) and standard IMRT (96.6%), despite experiencing consecutive feathering early in the treatment. **Figure 2g** depicts the larynx’s recovery.

**Figure 2:**
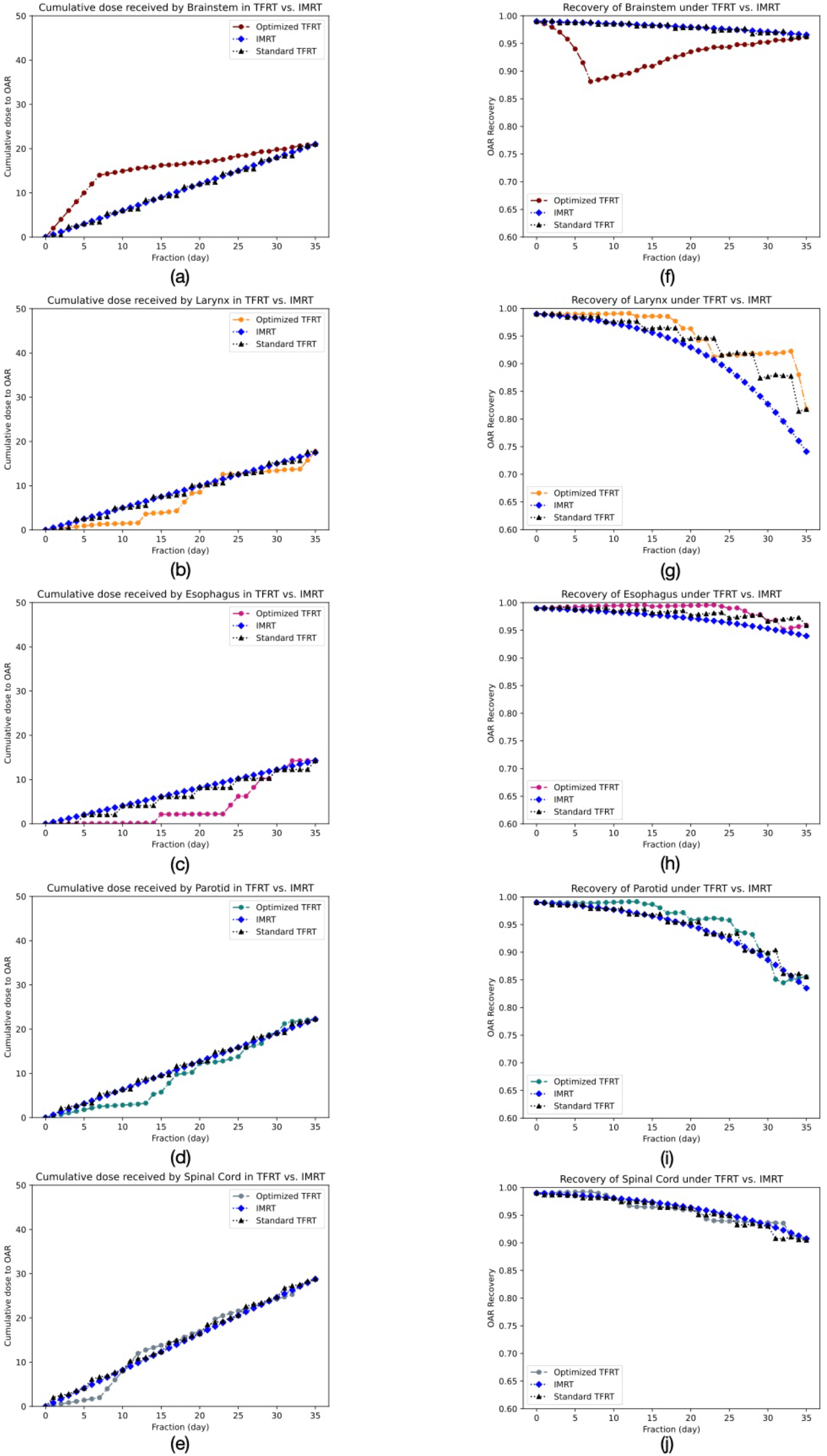
Comparison of Intensity Modulated Radiation Therapy (IMRT), standard Temporally Feathered Radiation Therapy (TFRT), and optimized dose-restricted TFRT when each Organ-at-Risk (OAR) has equal weights. Cumulative dose delivered to (a) brainstem, (b) larynx, (c) esophagus, (d) parotid, and (e) spinal cord are shown. Similarly, the recovery curves for (f) brainstem, (g) larynx, (h) esophagus, (i) parotid, and (j) spinal cord are shown.

Due to having the highest *α*/*β* ratio, the larynx suffers from significant damage on high-dose days and experiences slower recovery during rest days. However, the optimized TFRT, which allowed the larynx to rest for four and ten days between high-dose days, led to significantly improved recovery of 81.8% compared to standard IMRT (74.1%).

**Figure 2h** shows that the esophagus also benefited from TFRT. With the largest *β* value and an *α*/*β* ratio of around 3, it was initially expected to suffer considerable damage. However, due to its very low sparing factor of 0.005 on low-dose days, it experienced minimal radiation-induced damage, leading to an improved recovery of 95.9% compared to standard IMRT (93.9%). **Figure 2i** highlights that TFRT improved the recovery of the parotid gland relative to standard IMRT. Although the parotid has a *α*/*β* ratio of around 3 and relatively high *β*, its higher sparing factors meant it received greater doses on low-dose days compared to the brainstem, larynx, and esophagus. The variable rest days allowed the parotid to achieve better recovery (85.6%) by the end of the treatment compared to standard IMRT (83.5%). Finally, **Figure 2j** depicts the recovery of the spinal cord. With the lowest *α* and *β* values and a *α*/*β* of around 3, the spinal cord has the highest sparing factors among the OARs. Despite receiving higher doses on low-dose days, the resulting radiation-induced damage was lower due to its low *α* and *β* values, leading to recovery behavior (90.5%) for optimized TFRT which is similar to that observed with standard IMRT (90.8%). The recovery levels for each OAR at the end of treatment were almost identical under both standard TFRT and the optimized TFRT, as we ensured that the cumulative dose delivered to each OAR was equal in both approaches. This resulted in a treatment plan where each OAR was feathered exactly seven times and administered low doses 28 times under both standard TFRT and the optimized TFRT. As we considered fixed recovery rates throughout the treatment, the overall change in the recovery remained the same for each OAR. However, unlike standard TFRT, the optimized TFRT allowed variable recovery times between high-dose days.

### Dose-free TFRT with uniform recovery weights

As indicated in **Figure 3a**, the brainstem and spinal cord were feathered nine times under dose-free TFRT. The larynx esophagus, and parotid were feathered four, eight, and five times, respectively. Therefore, the brainstem, spinal cord, and esophagus were feathered more frequently when the cumulative dose constraints (Constraint (14)) were not considered in the model. The increased frequency of feathering for the brainstem and spinal cord can be attributed to their lower *α* values and higher recovery rates. On the other hand, the esophagus has higher *α* and *β* values and a lower recovery rate, and its low sparing factors likely contributed to its frequent feathering. Less frequent consecutive feathering compared to the initial experiment. While the end-of-the-treatment recovery for the brainstem (90.2%) and the spinal cord (87%) in this case was lower (**Figure 3b**) compared to the previous case (96.2% and 90.5%, respectively) (**Figure 1b**), the worst-case recovery (which occurred for the spinal cord) was improved compared to that for the dose-restricted TFRT (which occurred for larynx) (87% vs. 82%).

**Figure 3:**
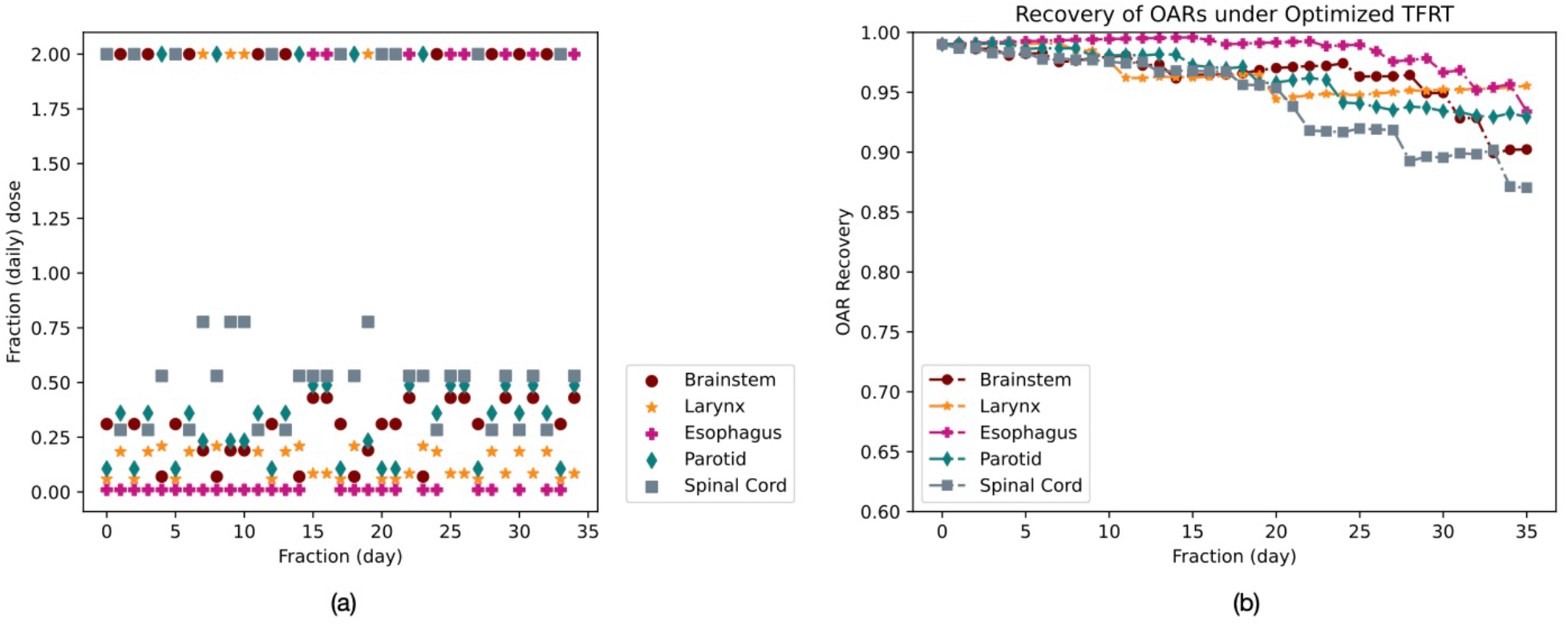
(a) The feathering schedule and (b) the recovery of OARs under optimized dose-free TFRT; OAR: organ-at-risk, TFRT: temporally feathered radiation therapy.

Again, due to its higher frequency of feathering, the brainstem received more doses under optimized TFRT (∼26 Gy) compared to standard IMRT (∼21 Gy) and standard

TFRT plans (∼21 Gy) (**Figure 4a**), resulting in poorer recovery (**Figure 4b**). The brainstem was feathered five times during the first half of the treatment, allowing it to recover by fraction 25, followed by four more feathering until the end of the treatment. **Figures 4c-4d** show cumulative dose received by the larynx and its recovery, respectively. The larynx was feathered the least, likely due to its higher *α* and *β* values, and a lower *α*/*β* ratio, which were accompanied by a slower recovery rate. Its infrequent feathering resulted in a lower cumulative dose and significantly better recovery (95.5%) by the end of treatment. Similarly, **Figures 4e-4f** present the recovery of the esophagus and the cumulative dose delivered. Despite being feathered more frequently and receiving a higher cumulative dose under optimized TFRT (∼16 Gy) compared to standard IMRT (∼14 Gy) and standard TFRT (∼14 Gy), its recovery by the end of treatment was comparable across all modalities, i.e., 93.4%, 94% and 95.9% for optimized TFRT, standard IMRT and standard TFRT, respectively. The esophagus was mainly feathered during the last 13 fractions, due to its very low sparing factors, which led to less radiation-induced damage. In other words, it was more advantageous to feather the OARs that sustained greater damage early in the treatment, allowing them additional time to recover before the treatment concluded.

**Figure 4:**
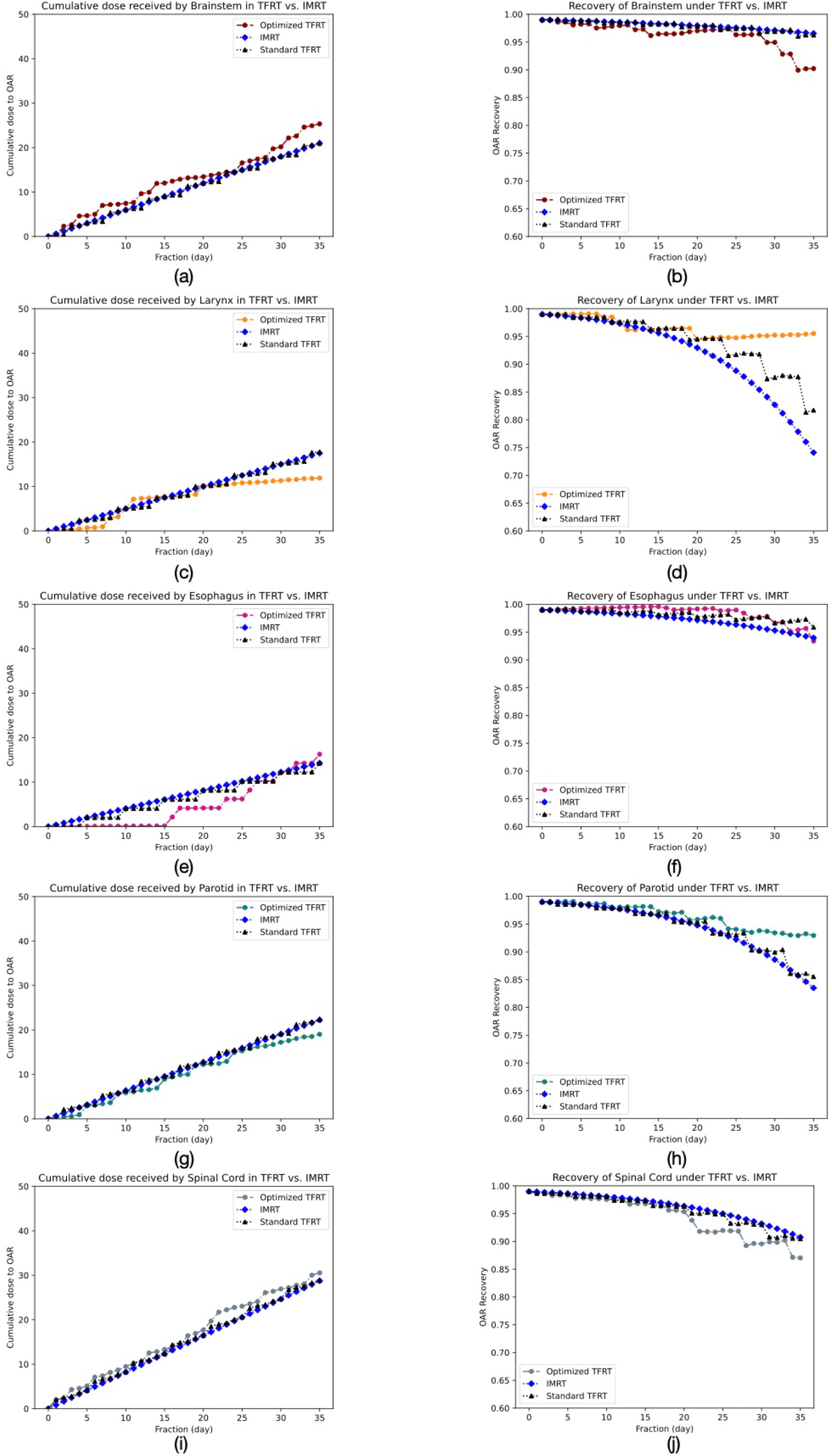
Comparison of Intensity Modulated Radiation Therapy (IMRT), standard Temporally Feathered Radiation Therapy (TFRT), and optimized dose-free TFRT when each Organ-at-Risk (OAR) has equal weights. (a) cumulative dose delivered to the brainstem (b) recovery of the brainstem (c) cumulative dose delivered to the larynx (d) recovery of the larynx (e) cumulative dose delivered to the esophagus (f) recovery of the esophagus (g) cumulative dose delivered to the parotid (h) recovery of the parotid (i) cumulative dose delivered to the spinal cord (j) recovery of the spinal cord.

**Figures 4g-4h** depict the recovery and cumulative dose received by the parotid. Feathered five times, the parotid received a lower cumulative dose of ∼19 Gy by the end of the treatment and showed significantly better recovery of 92.9%, as it was not feathered after fraction 25. **Figures 4i-4j** illustrate the recovery and cumulative dose delivered to the spinal cord. Despite being feathered nine times, the spinal cord received a slightly higher cumulative dose of ∼31 Gy than under standard IMRT (∼29 Gy) conditions. At the end of fraction 33, recovery levels were similar between standard TFRT (91%) and the optimized TFRT (90.2%), though the spinal cord had received more doses under the optimized TFRT. However, its recovery at the end of treatment was worse (87%) because the spinal cord was feathered at fraction 34.

### Dose-free TFRT with nonuniform recovery weights

For this analysis, we set the recovery weight of the spinal cord to 2, while maintaining a unit recovery weight for the rest of the OARs (**Supplementary Document A, Supplementary Figures S1a-S1b** and **Supplementary Figures S2a-S2j**). As a result, the spinal cord was feathered seven times which is less frequent compared to the uniform-weight cases (nine times), demonstrating the model’s capability to reflect the clinician’s preferences in prioritizing certain OAR’s recovery.

## Discussion

Mathematical modeling in radiation oncology allows for deconvolution of complex nonlinear biological mechanisms to generate and test hypotheses.^38,39^ By recognizing the variable radiosensitivity profiles across multiple OARs of HNC, our model implements TFRT with variable rest time for the OARs. The original TFRT study investigates a simple nonuniform spatiotemporal fractionation at the OAR level while maintaining uniform dose delivery to the tumor. Our model extends this concept by allowing more complex spatiotemporal alteration of the dose to the OARs. Unlike the original TFRT study (with a fixed 7-day rest time), our model leverages the LQ model within a dynamical NTCP model to guide the treatment in reducing the toxicity burden of certain OARs through, for example, early feathering of OARs with lower *α*/*β* ratios, providing them with more recovery time until the end of the treatment.

The biological heterogeneity in the tumor cells and across various OARs pose a significant challenge to the well-accepted biological assumptions on the effects of radiation therapy, leading to suboptimal oncological and toxicity outcomes.^38^ The OARs surrounding the tumor often react differently to radiation due to the variations in cellular repair mechanisms and each OAR’s intrinsic radiosensitivity. As indicated through the results of our optimized TFRT model, this complexity can allow for more tailored radiation delivery by considering organ’s unique radiosensitivity and recovery dynamics, potentially reducing toxicity and preserving the function of critical organs, which can lead to an overall improvement in patient quality of life during and after treatment.

In comparing the results of our model with those of the original TFRT, it is also important to note that the extra rest time for OARs in the original TFRT study may only become beneficial in certain cases depending on the amount of dose delivered to the OARs on their high- and low-dose days. While this indicates the potential benefit of the standard TFRT, it does not fully demonstrate the true potential of nonuniform spatiotemporal fractionation for the OARs, as the inherent radiosensitivity differences between the OARs are not incorporated into the analysis. In particular, our model demonstrated a notable improvement in the recovery of the spinal cord when nonuniform recovery weights were applied. Unlike the standard TFRT, which assumes uniform recovery time, our approach allowed the spinal cord to be feathered less frequently in the second half of the treatment, providing it with ample recovery time. As a result, the spinal cord achieved a slightly better recovery level compared to standard IMRT and original TFRT approaches, despite receiving a similar cumulative dose. This finding underscores the potential benefits of incorporating nonuniform recovery times into treatment planning, as it allows for more strategic dosing that aligns with each organ’s specific recovery characteristics. In contrast, our study allows a more comprehensive study of the potential outcomes of TFRT by deriving more complex treatment plans that recognize the nonhomogeneous radiosensitivity across multiple OARs.

The optimized TFRT model is also capable of deriving the optimal feathering schedule when preserving certain OARs that may be preferred over other OARs. For various reasons, the clinician may choose to prioritize certain OARs, e.g., parotid. For example, if the patient starts the treatment with some initial level of xerostomia, the clinician may choose to deliver the treatment plan that reduces the toxicity to the parotid more than what is expected under an IMRT treatment plan. As indicated in our results, our model can handle such priorities by varying the weights assigned to the recovery of the OARs by the end of the treatment. This flexibility in prioritizing organs based on individual patient needs could significantly enhance personalized treatment planning.

As explored in ART framework, shifting from uniform fractionation to delivering variable daily doses has been shown to improve tumor control and/or reduce normal tissue complication probabilities, particularly in treatments with frequent fractionation. Our model aligns closely with ART’s objectives by utilizing the variable daily dose received by the HNC OARs to enhance organ sparing, thereby reducing patient toxicity. Future research could explore incorporating daily on-treatment imaging data to further personalize the variable daily dose and optimize OAR recovery time, thus improving the quality of life of the HNC survivors.

Our model has limitations. However, the optimized TFRT model relies on estimations of *α* and *β* values and the optimal feathering schedule is highly dependent on the choice of radiosensitivity parameters. While the LQ model is established as a well-known dose-response prediction tool, the true values of the radiosensitivity parameters have been the subject of several studies with significant heterogeneity in the reported values.^18^ Similar ambiguity may arise when estimating other parameters in our NTCP dynamical model (e.g., the recovery rate *μ* and the recovery threshold *τ*). In the absence of any data to calibrate evolutionary dynamics, we have assumed these parameters to be constant during the course of radiation, which may be a significant simplification.

Further, one may discuss the choice of sparing factors to substitute the spatial distribution of the dose around OARs. To demonstrate the proof of concept, we have intentionally avoided the embedded dose-optimization task within the TFRT treatment planning. As a result, we have utilized the sparing factors to represent the possibly nonuniform spatial distribution of the dose received by OARs. However, further improvement in toxicity reduction may be achievable if the feathering schedule and dose optimization are simultaneously optimized. Future research could focus on integrating these aspects into a unified optimization model and validating its effectiveness in clinical settings.

## Conclusion

Temporally feathered radiation therapy (TFRT) for head-and-neck radiation therapy, when optimized based on the specific non-identical radiosensitivity profiles of OARs, can lead to better end-of-the-treatment recovery, thus offering the potential to improve patient toxicities. OAR-prioritized TFRT plans warrant further consideration in future radiation treatment planning for head-and-neck cancer patients as they reflect the clinician’s preference in certain OAR sparing.

## Data Availability

All data generated and analyzed during this study are included in this article (and its supplementary document). The source code is accessible through https://figshare.com/s/adc2470447ea492338c2.

## Supplementary Document: Dose-free TFRT with nonuniform recovery weights

Using our model, we also investigated the effect of reflecting the preferences of the clinician in prioritizing the recovery of certain OARs. For this analysis, we set the recovery weight of the spinal cord to 2, while maintaining a unit recovery weight for the rest of the OARs. **Figures S1a-S1b** present the prescribed doses to the OARs and their recovery outcomes when we do not restrict the cumulative dose delivered to each OAR and preserving the spinal cord is prioritized. This experiment suggested a treatment plan where the brainstem was feathered nine times. The spinal cord and esophagus were feathered seven times. The larynx and parotid were feathered six times. In contrast, the spinal cord was feathered less frequently compared to the uniform weight case, as the model prioritized its recovery towards the end of treatment. Specifically, the spinal cord was feathered six times in the first 13 fractions and once afterward, allowing sufficient time for recovery by the end of the treatment. The larynx, with its highest *α*/*β* ratio, did not benefit as much from lower fractionation doses compared to other OARs. Consequently, the model focused on feathering OARs with lower *α*/*β* ratios earlier in the treatment, while those with higher *α*/*β* ratios, such as the larynx, were feathered later.

**Figure S1:**
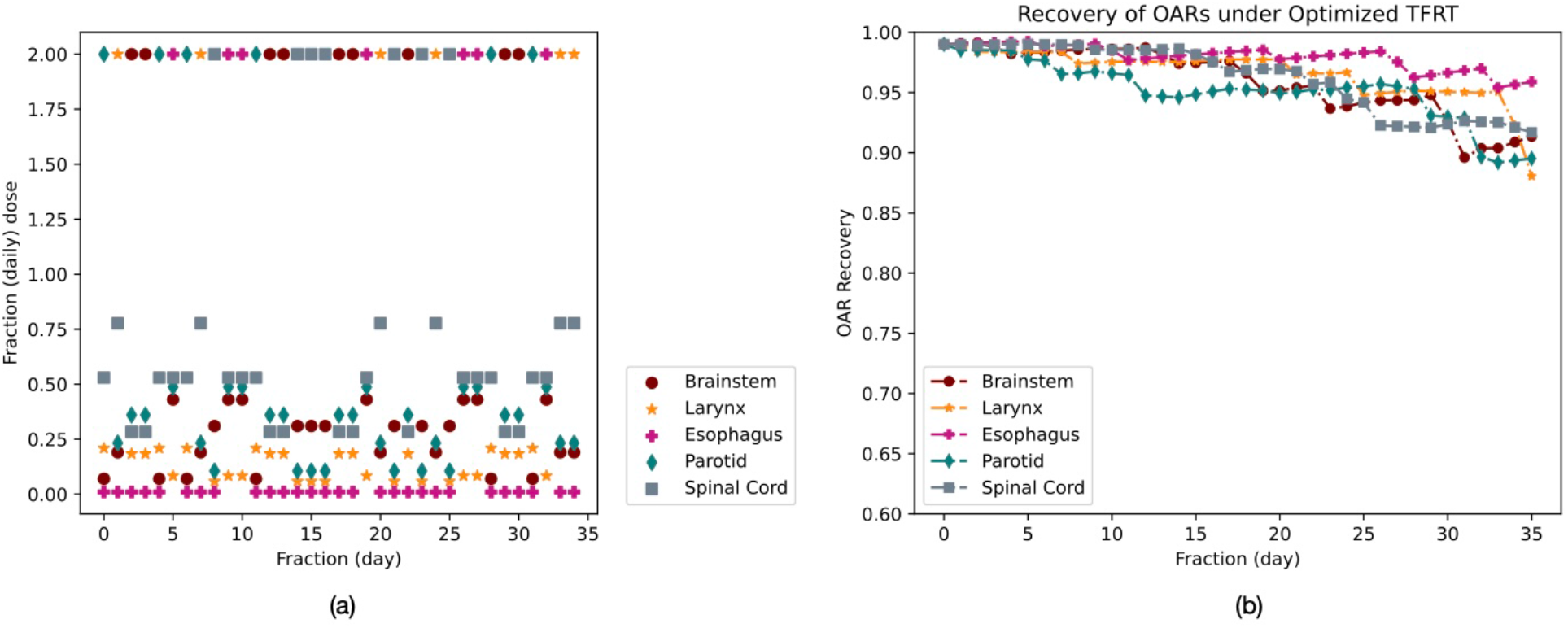
(a) The feathering schedule and (b) the recovery of OARs under optimized dose-free TFRT with nonuniform weights; OAR: organ-at-risk, TFRT: temporally feathered radiation therapy.

**Figures S2a-S2b** illustrate the recovery and cumulative dose delivered to the brainstem. Similar to the case of the uniform weight, the brainstem received more doses compared to standard IMRT and standard TFRT plans, leading to poorer recovery. By the end of fraction 28, the brainstem’s recovery (94.8%) was comparable to the recovery in the other treatment modalities (97.2% and 96.9% for standard IMRT and standard TFRT, respectively) despite higher cumulative doses. However, after being feathered three additional times post-fraction 28, its recovery level dropped noticeably, though it maintained a recovery level of 91% by the end of treatment. **Figures S2c-S2d** show the recovery and cumulative dose delivered to the larynx. The larynx was feathered more frequently compared to the uniform weight case but less than in the initial experiment. This resulted in a slightly lower cumulative dose and significantly better recovery by the end of treatment, i.e., 88.1%, 74.1% and 81.8% for optimized TFRT, standard IMRT and standard TFRT, respectively. **Figures S2e-S2f** present the recovery of the esophagus and the cumulative dose delivered. It was feathered less frequently than with uniform weights, with its cumulative dose remaining comparable to IMRT and standard TFRT under nonuniform weights. Consequently, its recovery (95.9%) at the end of treatment was better than IMRT (93.9%) but similar to standard TFRT (95.9%). **Figures S2g-S2h** depict the recovery and cumulative dose delivered to the parotid. The parotid was feathered more frequently compared to the case with uniform weights. It received a slightly lower cumulative dose (∼21 Gy) by the end of the treatment and showed significantly better recovery of 89.5% due to its spread-out feathering schedule while standard IMRT and standard TFRT exhibited recovery levels of 83.5% and 85.6%, respectively. **Figures S2i-S2j** illustrate the recovery and cumulative dose delivered to the spinal cord. Feathered seven times, the spinal cord received a cumulative dose nearly equivalent to that under standard IMRT conditions (∼29 Gy). By being feathered only once during the second half of the treatment, at fraction 20, and allowing a rest period for the remaining 15 fractions, the spinal cord achieved slightly better recovery (91.7%) compared to IMRT (90.8%) and standard TFRT (90.5%).

**Figure S2:**
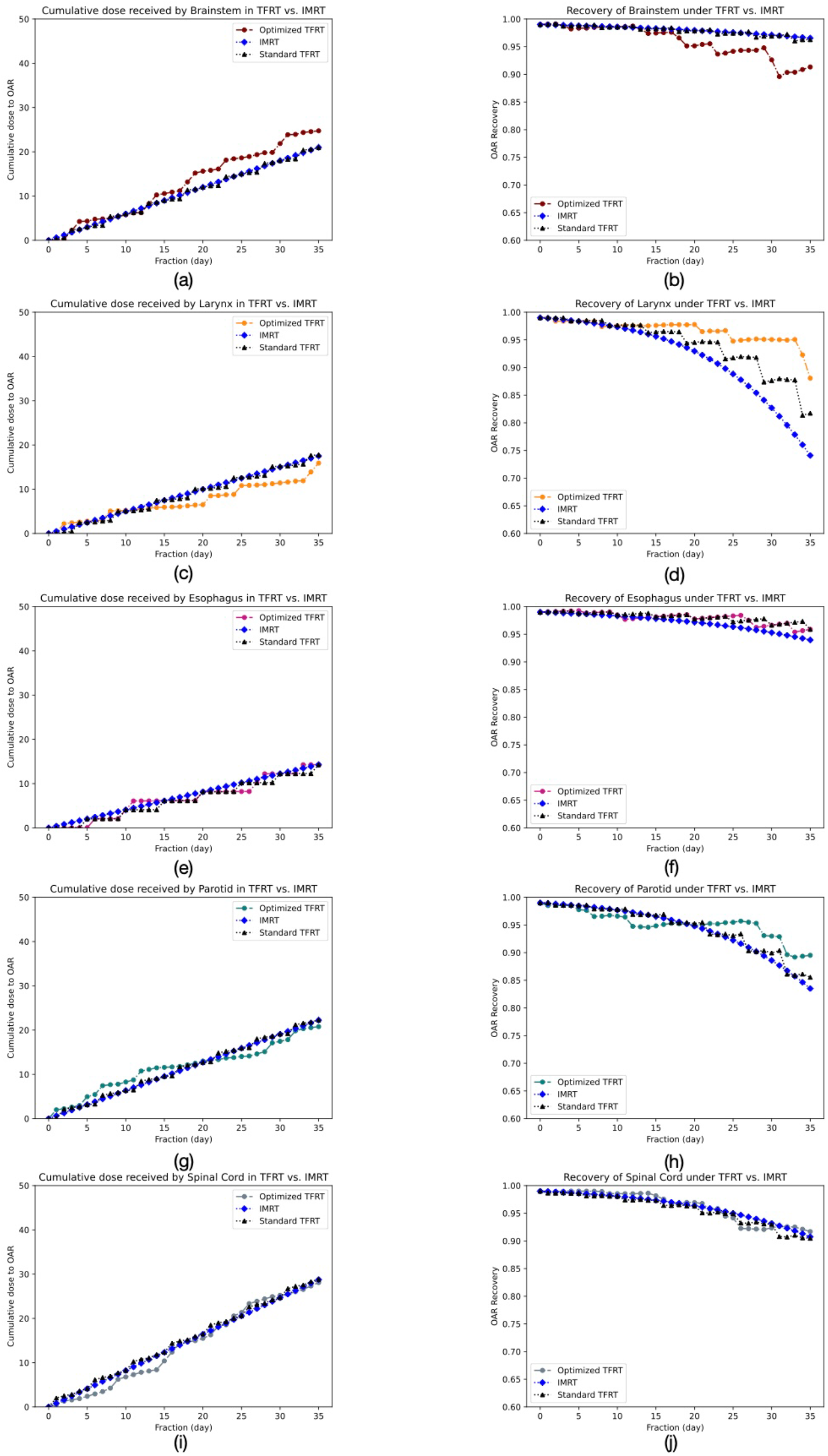
Comparison of Intensity Modulated Radiation Therapy (IMRT), standard Temporally Feathered Radiation Therapy (TFRT), and optimized dose-free TFRT when the spinal cord is prioritized twice other organs-at-risk (OARs). (a) cumulative dose delivered to the brainstem (b) recovery of the brainstem (c) cumulative dose delivered to the larynx (d) recovery of the larynx (e) cumulative dose delivered to the esophagus (f) recovery of the esophagus (g) cumulative dose delivered to the parotid (h) recovery of the parotid (i) cumulative dose delivered to the spinal cord (j) recovery of the spinal cord.

